# Artificial intelligence applications for dementia: A systematic review for clinical research

**DOI:** 10.1101/2025.08.22.25333992

**Authors:** Sergio Altares-López, Boris-Stephan Rauchmann

## Abstract

Artificial intelligence and emerging technologies are driving a transformative shift in society, particularly in the healthcare sector, where they enhance diagnostic accuracy, enable personalized treatment approaches, and improve patient monitoring. This systematic review presents a comprehensive analysis of current AI applications specifically emphasizing on dementia. We also address the primary challenges associated with the integration and widespread adoption of AI technologies, including issues related to data integration, model interpretability, regulatory barriers, and ethical concerns surrounding patient data privacy. Finally, we examine the future directions of AI in the diagnosis and treatment of dementia diseases, focusing on innovations in brain imaging, neuromodulation, and real-time monitoring technologies.

## 1. Introduction

Artificial intelligence is transforming medicine by improving disease detection, diagnosis, and treatment. With advancements in machine learning and data-driven analytics, it is becoming an essential tool in modern healthcare. The increasing integration of these technologies in medical practice suggests a growing trend toward more efficient, accurate, and personalized healthcare solutions.

An area where these models are increasingly being applied is dementia. Dementia refers to a group of progressive neurological disorders characterized primarily by cognitive decline, including memory impairment, reduced reasoning, and language difficulties. The integration of advanced modeling techniques in this domain supports earlier diagnosis, enables more accurate monitoring of disease progression, and facilitates the development of personalized treatment strategies. Dementia is not only devastating to those affected, but they also pose significant challenges to society. Currently, it is estimated that over 50 million people worldwide live with dementia, and this number is expected to double by 2050 due to the aging global population [102]. Dementia alone affects an estimated 5.8 million people in the United States, with similar numbers in Europe and other parts of the world. The global prevalence is rising rapidly, placing an immense burden on healthcare systems, families, and communities [69]. In addition to their impact, dementia contributes significantly to societal and economic challenges. The care of individuals with this syndrome often requires long-term support, with costs for healthcare services, medications, and caregiving services becoming financially overwhelming for families and governments [62]. In the United States, the annual cost of dementia care is estimated to be over $350 billion, and this figure is expected to rise exponentially as the population ages.

Traditional diagnostic and treatment approaches for dementia face significant limitations [59]. Current diagnostic methods for dementia often depend on clinical assessments and imaging techniques that detect the condition only after irreversible brain damage has occurred. Moreover, most available treatments aim to manage symptoms rather than target the underlying disease mechanisms. This highlights the urgent need for more effective, early-stage interventions, though recent advances, such as the disease-modifying drugs *Lecanemab*, offer a promising exception [97]. Given the complex and multifactorial nature of dementia diseases, there is an increasing demand for advanced technologies capable of improving early diagnosis, personalized treatment strategies, and continuous patient monitoring.

Artificial intelligence is transforming how we face dementia diseases by utilizing advanced machine learning, multimodal data integration, and predictive analytics. These algorithms can analyze vast amounts of heterogeneous data, including neuroimaging scans, genetic information, biomarkers, speech patterns, and wearable sensor data, to uncover disease-specific patterns that may not be apparent to human clinicians supporting precision medicine and personalized care. Furthermore, AI-enhanced drug discovery methods are accelerating the identification of potential therapeutic compounds, offering new hope for disease-modifying treatments. Despite its transformative potential, implementing AI in research and practical healthcare is challenging. Data privacy, model interpretability, regulatory approval, and ethical considerations must be carefully addressed to ensure the responsible and effective deployment of these solutions. Additionally, the need for large, high-quality datasets for training AI models presents logistical and technical hurdles.

This systematic review provides a comprehensive, overview of the role of artificial intelligence in dementia.

We explore the latest developments in AI-driven diagnostic tools, predictive modeling, and personalized treatment approaches while addressing key challenges and ethical considerations associated with AI integration in neurological care. Finally, we discuss future research directions and emerging trends that can further enhance AI’s impact in this domain, offering a roadmap for the continued evolution of AI applications in dementia disease management.

## 2. Systematic Review Criteria

A systematic literature search was conducted across PubMed, ACM, Springer, IEEE Scopus, Google Scholar, and Web of Science covering the period from January 2010 to March 2025. The purpose of the search was to identify studies applying artificial intelligence and machine learning methods in the context of dementia research and clinical care. The search strategy combined controlled vocabulary (e.g., MeSH terms) and free-text keywords related to both artificial intelligence and dementia.

Boolean operators were used to combine terms and ensure comprehensive coverage. Queries included variations such as *“artificial intelligence”, “machine learning”, “deep learning”, “neural network”, “transformer”, “natural language processing”, “LLMs”, “generative models”*, combined with clinical concepts like *“dementia”, “Alzheimer”, “frontotemporal dementia”, “neurodegenerative”*, and *“vascular dementia”*, as well as outcome-related descriptors such as *“diagnosis”, “classification”, “prediction”, “biomarker”*, and *“monitoring”*. The complete list of terms is presented in Table 1. Additionally, reference lists of included studies and relevant reviews were screened manually to identify studies not retrieved through electronic searches.

**Table 1.** Literature Search Strategy.

| Element | Details |
| --- | --- |
| Databases | PubMed, ACM, Springer, IEEE, Scopus, Google Scholar, Web of Science |
| Period | January 2010 – March 2025 |
| Objective | Identify studies applying artificial intelligence (AI) and machine learning (ML) methods in dementia research and clinical care |
| Search Strategy | Combination of controlled vocabulary (e.g., MeSH terms) and free-text keywords related to AI and dementia |
| AI-related Terms | *artificial intelligence, machine learning, deep learning, neural network, transformer, natural language processing, LLMs, AutoGPT, CNN, RNN, generative models, XAI, interpretability, Shap, LIME, self-supervised learning, SSL, Grad-CAM, ethics, Agentic AI, SVM, Random forest* |
| Clinical Terms | *dementia, Alzheimer, frontotemporal dementia, neurodegenerative, vascular dementia* |
| Outcome-related Terms | *diagnosis, multimodal, classification, prediction, biomarker, neuroimaging, MRI, repurposing, EEG, APOE, MEG, drug discovery, monitoring, EHR, regulations, data protection* |
| Terms Combination | AND, OR used to combine terms and ensure comprehensive coverage |
| Manual Screening | Reference lists of included papers and relevant reviews were screened to capture additional studies not retrieved electronically. Preprints have been excluded except those with a sufficient number of citations with applications in the field. Also, the current regulations and laws are considered. |

### 2.1. Eligibility Criteria

Eligibility criteria were defined a priori according to PRISMA 2020 guidelines. Original empirical research published in peer-reviewed journals or conference proceedings applying AI/ML to human subjects or real-world clinical datasets in dementia was included. Studies addressing diagnostic classification, prognostic modeling, risk stratification, biomarker discovery, clinical decision support, or multimodal data integration, and published in English between January 2010 and March 2025, were eligible.

We excluded studies based on simulated or synthetic data without clinical correlation, non-human or preclinical models, narrative reviews, editorials, commentaries, opinion pieces, publications lacking methodological transparency, and non-indexed or non–peer-reviewed sources (see Table 2). Titles and abstracts of all unique records were screened according to these criteria, and potentially relevant articles were assessed in full text. Disagreements were resolved by discussion and consensus. The final set of included studies constituted the evidence base for this review, summarized in the PRISMA 2020 flow diagram (Figure 1).

**Table 2.** Inclusion and exclusion criteria applied in the systematic review.

| Category | Inclusion Criteria | Exclusion Criteria |
| --- | --- | --- |
| Population | Human participants, real-world clinical datasets | Animal studies, simulated or synthetic data only |
| Study Type | Original peer-reviewed research (journal or conference) | Reviews, commentaries, editorials, opinion pieces |
| Language | English | Non-English publications |
| Time Frame | Published between Jan 2010 – Mar 2025 | Outside this range |
| Scope | AI/ML applied to dementia: diagnosis, prognosis, biomarker discovery, monitoring | Studies not involving dementia or not applying AI/ML |
| Data | Clinical, neuroimaging, biomarker, text, EEG/MEG, multimodal | Insufficient methodological transparency (e.g., unclear data, model, or metrics) |
| Peer Review | Indexed journals or recognized conference proceedings | Non-indexed sources, no peer-review process |

**Table 3.** Clinically relevant MRI sequences, AI tasks, and their role in dementia disease diagnosis. (*) CSF: Cerebrospinal Fluid.

| MRI Sequence | Key Features | Importance in dementia diseases | AI Tasks |
| --- | --- | --- | --- |
| **T1-weighted** | Provides contrast for visualizing structural anatomy; grey and white matter stand out, *CSF appears dark. | Detects structural changes and brain atrophy. | Image segmentation, anomaly detection, pattern recognition. |
| **T2-weighted** | Highlights edema and tissue damage; fluids like *CSF appear bright. | Detects tissue alterations and abnormalities. | Lesion detection, image classification, feature extraction. |
| **FLAIR (Fluid Attenuated Inversion Recovery)** | Suppresses *CSF signals, improves lesion visualization in periventricular and fluid-filled regions. | Enhances detection of abnormalities in white matter. | Lesion segmentation, progression tracking, anomaly detection. |
| **T2\*** | Sensitive to magnetic susceptibility; identifies micro-hemorrhages or iron deposits. | Helps assess iron accumulation and small hemorrhages. | Signal variation detection, image enhancement, anomaly detection. |
| **Diffusion-weighted Imaging (DWI)** | Assesses water molecule movement in brain tissue, providing insights into microstructural changes. | Useful for detecting early microstructural alterations. | Diffusion pattern analysis, image enhancement, structural analysis. |

**Table 4.**
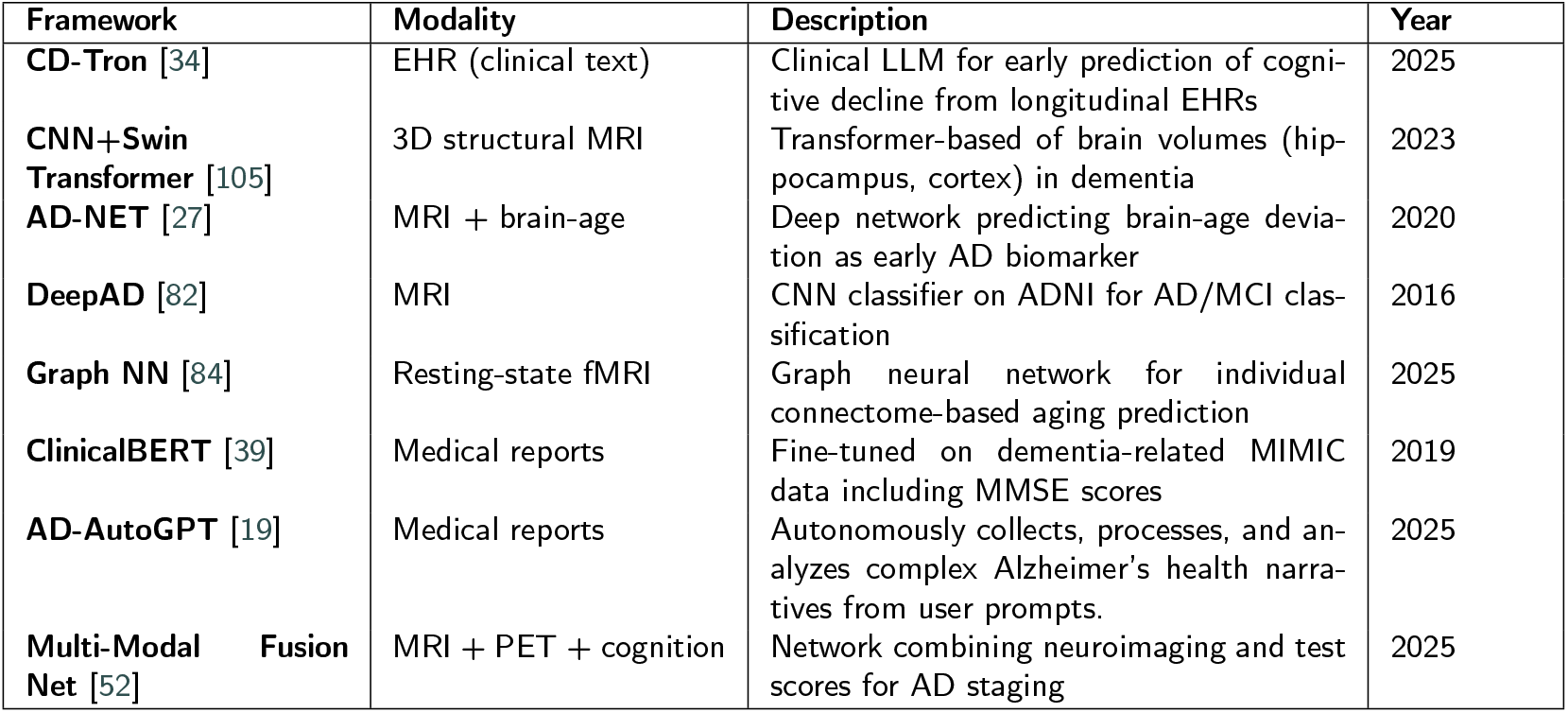
Representative AI models applied to dementia-related data across modalities.

**Table 5.**
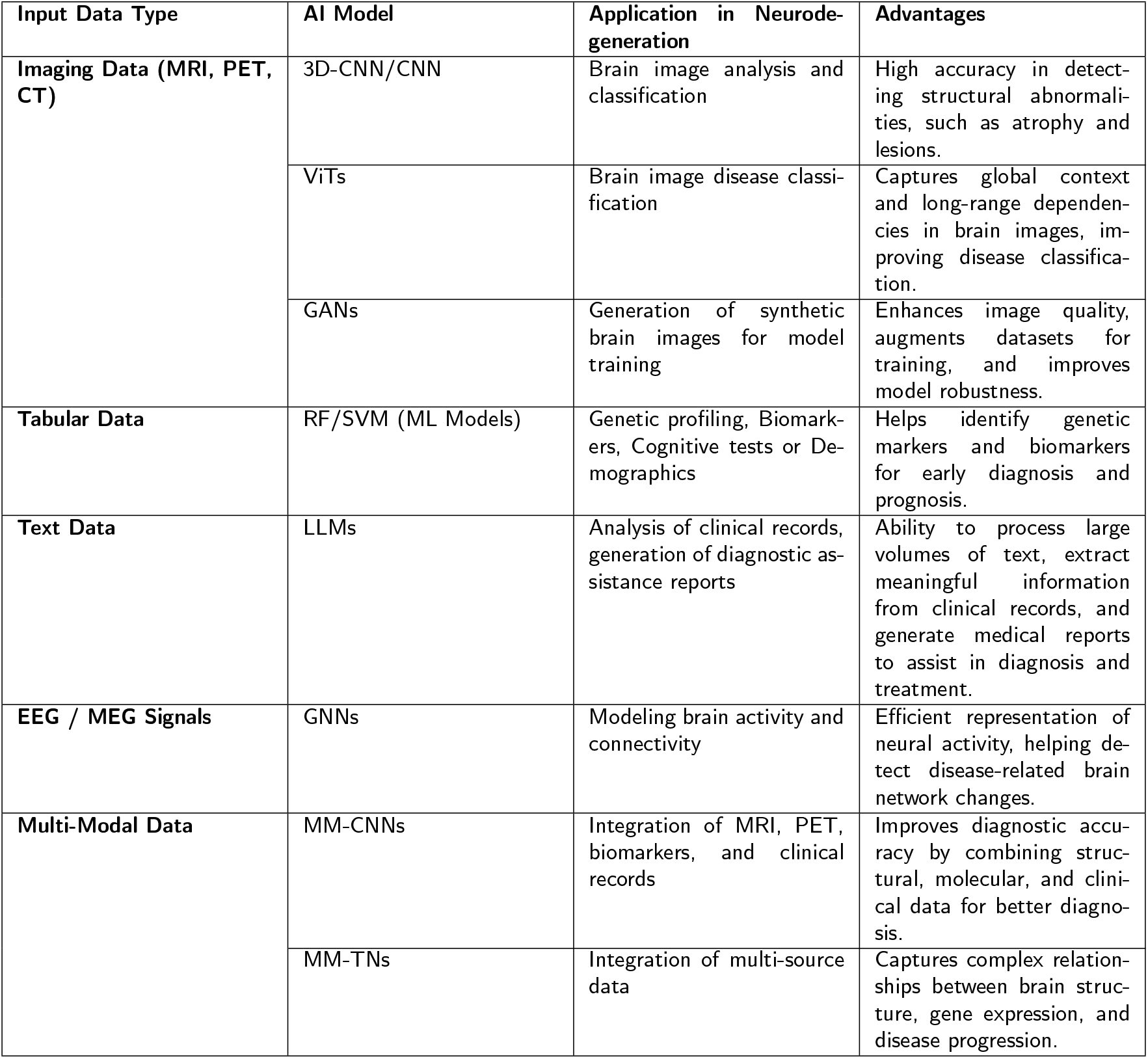
AI Models in Neuroscience Applications by Input Data Type.

| Input Data Type | AI Model | Application in Neurodegeneration | Advantages |
| --- | --- | --- | --- |
| Imaging Data (MRI, PET, CT) | 3D-CNN/CNN | Brain image analysis and classification | High accuracy in detecting structural abnormalities, such as atrophy and lesions. |
|  | ViTs | Brain image disease classification | Captures global context and long-range dependencies in brain images, improving disease classification. |
|  | GANs | Generation of synthetic brain images for model training | Enhances image quality, augments datasets for training, and improves model robustness. |
| Tabular Data | RF/SVM (ML Models) | Genetic profiling, Biomarkers, Cognitive tests or Demographics | Helps identify genetic markers and biomarkers for early diagnosis and prognosis. |
| Text Data | LLMs | Analysis of clinical records, generation of diagnostic assistance reports | Ability to process large volumes of text, extract meaningful information from clinical records, and generate medical reports to assist in diagnosis and treatment. |
| EEG / MEG Signals | GNNs | Modeling brain activity and connectivity | Efficient representation of neural activity, helping detect disease-related brain network changes. |
| Multi-Modal Data | MM-CNNs | Integration of MRI, PET, biomarkers, and clinical records | Improves diagnostic accuracy by combining structural, molecular, and clinical data for better diagnosis. |
|  | MM-TNs | Integration of multi-source data | Captures complex relationships between brain structure, gene expression, and disease progression. |

**Figure 1:**
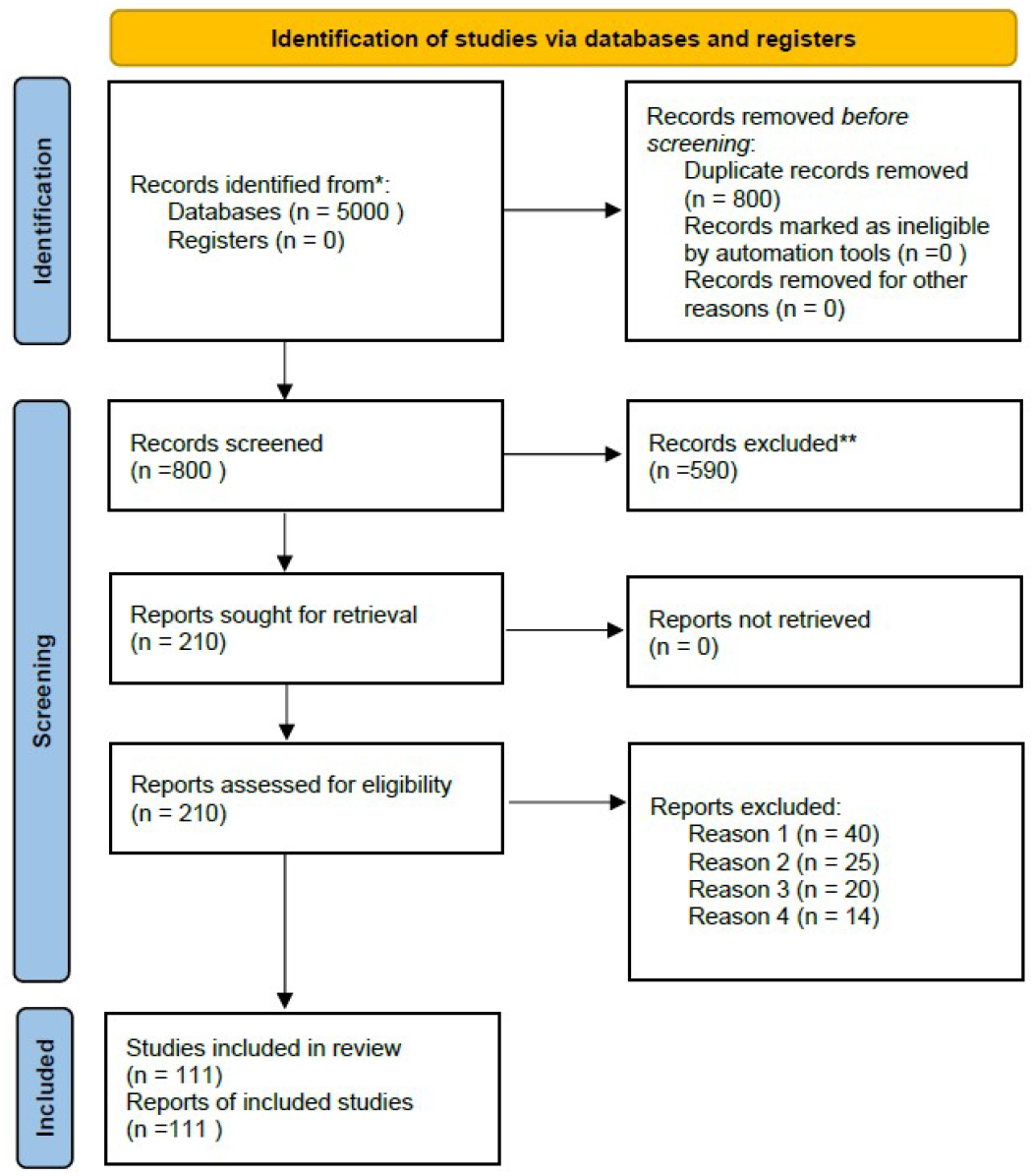
PRISMA 2020 Multistage screening and eligibility assessment process for the systematic review of AI applications in dementia research.

### 2.2. Data Extraction

To ensure consistency and transparency, data were extracted using a structured data extraction sheet that was developed prior to the review. For each included study, we recorded bibliographic details such as authors, year, country of origin, and publication source, together with information on study design and dataset characteristics, including sample size, clinical setting, and data modality. We also extracted detailed descriptions of the artificial intelligence and machine learning methods applied, including the type of algorithm or model, the strategy used for training and validation, and the reported performance metrics. In addition, we documented the input modality (such as neuroimaging, tabular data, clinical biomarkers, text, multimodal data, or EEG/MEG), the specific clinical application targeted (diagnosis, prognosis, biomarker identification, monitoring, or treatment support), and the main findings.

### 2.3. Potential Bias and Limitations

Although this systematic review followed PRISMA 2020 guidelines to ensure rigor and reproducibility, several potential sources of bias should be taken into account. First, publication bias may be present, as studies with positive or significant findings are more likely to be published, while negative or null results may remain underreported. Second, the restriction to English-language publications could introduce language bias, excluding relevant evidence from non-English sources. Third, despite the comprehensive search strategy across multiple databases, the fast-paced nature of artificial intelligence research means that some recent or unpublished works (e.g., preprints, conference proceedings without peer review) might not have been captured. Furthermore, heterogeneity between study designs, datasets, and AI methods complicates direct comparison and may limit the generalizability of the findings. Finally, the absence of a formal risk of bias assessment for each included study, represents another limitation.

## 3. Artificial Intelligence Applications for the Diagnosis and Stratification of Dementia

Artificial intelligence has emerged as a powerful tool in studying and diagnosing dementia, offering advanced analytical capabilities across various types of medical data. By leveraging AI-driven approaches, clinicians can improve early detection, enhance diagnostic accuracy, and uncover novel insights into disease progression. This section examines the various applications of AI in dementia research, categorized by the type of data utilized. Specifically, we examine models applied to imaging, tabular data (such as biomarkers, cognitive test results, or clinical scores), textual information, and multimodal datasets, highlighting their impact on clinical practice.

### 3.1. Medical Imaging

Medical imaging has fundamentally advanced the diagnosis and monitoring of dementia by providing detailed, non-invasive visualization of brain structure and function. Magnetic Resonance Imaging (MRI), Positron Emission Tomography (PET), and Computed Tomography (CT) are routinely employed to detect early pathological changes, monitor progression, and assess treatment efficacy. MRI provides high-resolution anatomical images, which are essential for identifying structural abnormalities, while PET evaluates functional processes, including glucose metabolism and the deposition of dementia-related biomarkers, such as amyloid-beta and tau. CT, though less sensitive for soft tissues, is valuable for detecting gross structural changes, particularly when MRI is contraindicated. The integration of these modalities with artificial intelligence enhances diagnostic accuracy and efficiency by managing the complexity and volume of imaging data.

Within artificial intelligence, deep learning models have revolutionized the field of medical imaging by enabling automated and more accurate interpretation of complex datasets. Convolutional neural networks (CNN), which have been widely adopted in this domain, are designed to recognize patterns in image data by applying layers of convolutional filters. These networks have achieved remarkable success in various medical imaging tasks, such as tumor detection, organ segmentation, and disease classification [80, 23]. However, while CNN have contributed significantly to the field, more advanced architectures are now emerging to address their limitations, enabling even more precise analysis. One notable advancement is the use of Vision Transformers (ViTs), which are based on transformer architectures.

These models require vast quantities of data for training to achieve optimal performance. ViTs have shown superior performance in various image classification tasks, particularly in situations where large datasets are available. Unlike CNNs, which rely on localized convolution operations to capture spatial hierarchies in images, Vision Transformers operate by splitting the image into non-overlapping patches and processing them as a sequence, similar to how words are processed in Natural Language Processing (NLP) models [106, 35]. This approach enables ViTs to capture long-range dependencies and global context within images, which can be particularly valuable in understanding the widespread, complex atrophy patterns associated with dementia [83, 44]. In addition to ViTs, Generative Adversarial Networks (GANs) have also gained traction in medical imaging. These models consist of two neural networks—a generator and a discriminator—that work in opposition to improve the generation of realistic synthetic data brain scans, particularly when real clinical data is scarce or difficult to obtain [88, 79, 67]. GANs can also aid in improving image quality by reducing noise, enhancing resolution, and generating high-quality images from lower-resolution scans, which is particularly valuable in clinical settings with limited resources [10, 111].

Another cutting-edge technique gaining attention is Attention Mechanisms, including the Self-Attention used in transformer-based models. Attention mechanisms allow models to focus on the relevant features of an image while ignoring less important ones. These models focus on key regions of the brain that show early signs of neurodegeneration, improving diagnostic accuracy and reducing the risk of overlooking subtle changes [61, 101]. This approach is particularly beneficial in diseases where early-stage changes in specific brain regions, like the hippocampus, need to be detected for timely intervention.

Moreover, the integration of Multimodal Learning techniques is becoming increasingly important. This type of model combines multiple types of imaging data, such as different MRI modalities or PET scans, which have the potential to provide a more comprehensive understanding of disease pathology. Multi-Modal Convolutional Neural Networks (MM-CNNs) and other hybrid architectures, which integrate data from different imaging modalities, improve diagnostic performance by leveraging complementary information [90, 40]. By providing detailed structural information about brain anatomy, MRI complements PET, which reveals functional changes such as glucose metabolism and amyloid-beta or tau deposition, and their combination enables more accurate detection of both structural and functional abnormalities, leading to earlier and more precise diagnoses.

Advancements in 3D Imaging and Volumetric Analysis have enhanced the capability of AI models to assess the brain in three dimensions, providing more detailed and accurate information about the size, shape, and volume of affected regions. 3D CNNs are particularly suited for analyzing volumetric data as they are capable of capturing spatial relationships across three-dimensional brain scans. These models are used to assess brain atrophy patterns, which are important for tracking disease progression [15]. In addition, some models are also being developed to assist in predicting disease progression. Longitudinal studies that track the development of dementia diseases over time generate vast amounts of data, which machine learning models leverage to predict future disease trajectories. Recurrent Neural Networks (RNNs) and Long Short-Term Memory (LSTM) networks, both of which are designed to handle sequential data or time series, are particularly well-suited for modeling disease progression in these settings. These models can analyze changes in imaging data over time to predict the rate of cognitive decline, the onset of symptoms, and the expected response to treatment, enabling more personalized and proactive care [17, 56, 25].

### 3.2. Text Data and Language Models

In the clinical setting, vast amounts of unstructured text data are generated daily, including medical records, physician notes, and patient reports. Extracting meaningful insights from this information is crucial for improving diagnostics, treatment planning, and disease monitoring. Recent advances in natural language processing (NLP) and large language models (LLMs) have enabled more efficient analysis of these textual datasets, allowing for automated information retrieval, predictive modeling, and enhanced decision support systems. NLP techniques have proven to be an effective tool for the analysis of patients’ speech and writing [94]. Traditionally, NLP approaches have used models such as RNNs and LSTMs to detect language syntax, semantics, and fluency patterns that may indicate cognitive impairment. However, recent advances in Large-Scale Language Models (LLMs), such as GPT (Generative Pre-trained Transformer), BERT (Bidirectional Encoder Representations from Transformers), and specialized variants in the biomedical domain, have significantly expanded NLP’s capabilities in this field.

LLMs can process large volumes of clinical and speech data, identifying subtle correlations that may not be evident through conventional methods. Recent studies have shown that models such as GPT-4 can analyze transcripts of medical interviews and detect linguistic anomalies associated with dementia’s, such as word repetition, decreased grammatical complexity, and the use of generic terms instead of specific vocabulary [78]. Moreover, the integration of contextual embeddings enables these models to evaluate disease progression over time by performing longitudinal analyses of patients’ speech and writing samples and extracting relevant information from unstructured medical records.

Another important advance is the integration of these LLM techniques with multimodal diagnostic systems (See Section 3.4), where these models can combine linguistic information with neuroimaging data, biomarkers, and electronic medical records to provide a more accurate and personalised assessment of the patient’s cognitive status [24, 14]. In this context, models pre-trained on large biomedical datasets, such as BioBERT or ClinicalBERT, have shown significant improvements in extracting relevant clinical information and predicting the progression of dementia diseases [16, 49].

However, the use of LLMs in clinical applications and neuroscience also presents significant challenges. Interpretability remains a key problem, as models can generate predictions without clear justification, making their adoption in medical settings difficult. In addition, inherent biases in training data can affect diagnostic accuracy, especially in underrepresented populations. Furthermore, language and dialectal challenges persist, as most large models are predominantly trained on English data. This leads to reduced performance and reliability when applied to clinical contexts involving local languages, regional dialects, or multilingual patient populations, potentially exacerbating existing healthcare disparities. To mitigate these problems, explainable AI (XAI) approaches (See Section 5.2) and fine-tuning methods are developed with clinical data, which could improve the transparency and reliability of these models in real-world medical practice [28, 46].

### 3.3. Tabular Data: Biomarkers, Genetic, and Demographic

Structured clinical data plays a fundamental role in understanding and predicting dementia diseases. Unlike unstructured text or imaging data, tabular datasets integrate diverse yet complementary sources of information, including genetic predisposition, biochemical and molecular biomarkers, clinical evaluations, cognitive test scores, demographic variables, lifestyle variables, and genetic data such as the existence of the APOE gene [99, 29]. Machine learning models have been widely applied to identify risk factors associated with dementia diseases. Random Forest (RF) and Support Vector Machines (SVMs) are widely used to analyze these complex interactions between genetic variants, polygenic risk scores, and disease susceptibility [21, 89]. Deep learning approaches, such as autoencoders, have also been employed to reduce the dimensionality of large genomic datasets and extract meaningful representations for dementia prediction [72].

These models are widely used to analyze biochemical and molecular biomarkers, such as amyloid-beta plaques, tau tangles, neuroinflammation markers, and metabolic indicators derived from cerebrospinal fluid (CSF) and blood tests [9, 58]. Deep Neural Networks (DNNs) and convolutional neural networks have been employed to identify patterns in biochemical assays, mass spectrometry data, and other molecular profiling techniques. RNNs have also been leveraged to track biomarker evolution over time, enabling the prediction of disease progression [100].

On the other hand, clinical data —including cognitive test results, neuropsychological assessments, and medical history which usually are unstructured data— provide additional layers of information. Decision Trees and Gradient Boosting Machines (GBMs) have been applied to structured clinical datasets to classify disease stages and predict cognitive decline [26, 110]. Demographic and lifestyle factors, such as age, sex, education level, socioeconomic status, diet, physical activity, and sleep patterns, significantly influence dementia disease risk [70, 5]. Algorithms incorporating these features enhance predictive accuracy by capturing environmental and behavioral contributions to disease onset. Logistic regression, survival analysis models, and Bayesian networks have been also used to quantify these relationships and assess individual risk profiles [85].

Recently, transformer-based models have been introduced to enhance the analysis of heterogeneous tabular data.

The Transformer-based Prior-Data Fitted Network (TabPFN) has emerged as a state-of-the-art method for classifying small datasets without requiring extensive hyperparameter [37]. TabPFN has proven useful in the dementia domain when combined with LightGBM to predict conversion in Parkinson’s disease patients. The model achieved an accuracy of 95.92% and an AUC of 0.9737, outperforming seven existing methods [95]. Studies highlight its potential in distinguishing pathological from healthy patient profiles, offering a promising tool for early diagnosis and personalized risk assessment [91].

### 3.4. Multimodal AI Approaches

The integration of diverse data sources, such as medical imaging, biomarker data, and clinical information, has been shown to significantly improve the diagnostic accuracy of AI-driven systems. Multimodal AI approaches enable the fusion of different information streams, facilitating a more comprehensive understanding of disease processes. AI models that incorporate various data types, including imaging, genetic markers, and clinical data, provide a more complete and nuanced view of dementia diseases [73, 98]. Multi-Modal Convolutional Neural Networks (MM-CNNs) and Multi-Modal Transformer Networks (MM-TNs) combine imaging data (e.g., MRI, PET scans) with additional modalities, such as genetic and clinical data. These models enhance diagnostic accuracy by leveraging the complementary information each modality offers [60, 11]. The integration of MRI scans to detect structural changes with PET scans to capture molecular and functional alterations delivers a more thorough and accurate representation of the disease’s pathology.

Time-series data is critical for predicting the progression of dementia diseases. Recurrent Neural Networks (RNNs) and Long Short-Term Memory (LSTM) models are commonly employed to analyze longitudinal data, such as repeated cognitive assessments and brain imaging [36, 32]. These models predict the course of these diseases, enabling more informed interventions and management strategies [38]. Furthermore, Graph Neural Networks (GNNs), which are effective at modeling relational data, have been used to explore how the progression in one region of the brain’s network can influence the development of other regions. AI models, particularly those utilizing Random Forests (RF) and Gradient Boosting Machines (GBMs), have demonstrated success in predicting disease trajectories by analyzing tabular data. By examining data across multiple time points, these models predict the rate of cognitive decline, identify risk factors, and guide treatment strategies. These predictive capabilities are pivotal for personalized medicine, allowing proactive care and timely interventions for patients at risk of substantial cognitive decline. Recent advances, such as multimodal compound AI and multi-agent AI systems, further enhance this potential by integrating diverse data types (e.g., imaging, genetics, clinical history) and enabling collaborative decision-making among specialized AI agents. These systems improve robustness, interpretability, and adaptability in complex clinical scenarios, making them particularly suitable for neurodegenerative disease management [47, 107].

### 3.5. EEG and MEG Analysis

Electroencephalography (EEG) and Magnetoencephalography (MEG) are two neuroimaging techniques that play a crucial role in monitoring changes in neural activity in dementia diseases. EEG measures electrical activity in the brain through electrodes placed on the scalp, while MEG captures the magnetic fields produced by neuronal activity, offering higher spatial resolution.

EEG is widely used due to its non-invasive nature, affordability, and high temporal resolution, making it a practical tool for clinical and research applications [41]. AI enhances EEG analysis by applying advanced signal processing techniques and deep learning models to detect subtle changes in brainwave activity, serving as early indicators of cognitive decline, neuronal dysfunction, or disease progression. AI models analyze variations in EEG frequency bands—such as alpha, beta, delta, and theta waves—associated with conditions like dementia’s disease and other forms of dementia. Machine learning algorithms are increasingly employed to extract features from EEG signals, enabling the detection of deviations in brain network connectivity, spectral power, and event-related potentials (ERPs). These deviations serve as quantifiable markers for neurodegeneration [75, 50]. Furthermore, emerging methods based on quantum computing paradigms, the so-called quantum machine learning (QML), are beginning to explore these pathways [53].

MEG complements EEG by providing a more direct and precise representation of cortical activity, as it is not affected by the conductivity properties of the skull and scalp. AI-driven MEG analysis enables the detection of microscale alterations in neuronal oscillations and network connectivity, which may serve as early biomarkers for dementia diseases. Machine learning models can process MEG signals to differentiate between normal and pathological brain states, identifying disruptions in oscillatory dynamics and functional connectivity. One of MEG’s main advantages is its ability to pinpoint specific cortical and subcortical regions affected by neurodegeneration with high accuracy.

Researchers develop robust, multimodal biomarkers for early-stage diagnosis and disease progression tracking by correlating cognitive states derived from these techniques. EEG is particularly valuable in sleep analysis, as disruptions in sleep architecture are strongly linked to dementia diseases. AI models have shown to identify anomalies in REM and non-REM sleep patterns, offering critical insights into disease development. Meanwhile, MEG’s superior spatial resolution contributes to more precise biomarker discovery. The integration of AI-driven EEG and MEG analyses might enhance dementia disease assessments, ultimately supporting early intervention strategies and improving patient outcomes.

## 4. Treatment and Intervention Strategies

Artificial intelligence has emerged as a transformative tool in the development of strategies to treat and manage dementia diseases. In addition to improving the accuracy of early diagnosis, it allows for personalised treatments and accelerates the discovery of new drugs, thus optimising both research processes and patient care.

### 4.1. Drug Discovery and Personalized Medicine

The process of drug discovery is critical in the fight against dementia diseases, yet it has traditionally been a slow, expensive, and uncertain endeavor. These disorders, characterized by progressive neurological decline, have long posed significant challenges for researchers and clinicians, with many treatments failing to produce the desired outcomes. The rise of artificial intelligence is revolutionizing the field by streamlining drug development processes and offering innovative ways to identify promising drug candidates more quickly. AI is also opening new doors for repurposing existing treatments, which could significantly enhance the speed and success rate of discovering effective therapies. However, the rise of artificial intelligence is revolutionizing the field by streamlining drug development processes and offering innovative ways to identify promising drug candidates more quickly.

One of the most important advances in this field is the *virtual screening* and *molecular docking*. Virtual screening refers to the computational process of evaluating large libraries of chemical compounds to identify potential drug candidates that may bind to a target protein associated with a specific disease. This technique allows researchers to prioritize compounds for further experimental testing, significantly reducing the time and cost of drug discovery. On the other hand, molecular docking is a method used to predict the binding interaction between a small molecule (such as a potential drug) and a target protein. By simulating how these molecules fit together, molecular docking helps to assess the strength and specificity of the binding, providing valuable insights into the effectiveness of a drug candidate. These methods allow machine learning algorithms to simulate the interaction between pharmacological compounds and target molecules, such as specific proteins or receptors in the brain, without the need for physical trials [55, 74, 57]. Through computational prediction, it is possible to model how different molecules bind to disease-related proteins, allowing thousands of compounds to be filtered before laboratory testing. This approach greatly reduces the costs associated with the initial stages of drug discovery, accelerating the identification of promising compounds. Artificial intelligence also plays a crucial role in optimizing clinical trials, helping to accelerate the transition from laboratory discoveries to their clinical application. Algorithms analyze large volumes of previous clinical data, making it possible to identify the most effective and safest drugs for each type of patient. In addition, artificial intelligence optimizes the patient selection process, identifying those who meet the specific criteria of the clinical trial, thus ensuring that the studies are carried out with a representative and homogeneous group of people. This approach not only improves the quality of clinical trials, but also increases their effectiveness, allowing treatments to reach the market more quickly [104, 30].

In addition, artificial intelligence is facilitating the reuse of existing drugs, a process known as *repurposing* [76, 87].

This strategy seeks to identify new therapeutic indications for drugs that have already been approved for other diseases, thus bypassing early-stage development and safety testing [20]. AI contributes to drug repurposing by integrating and analyzing heterogeneous biomedical data sources—such as molecular structures, target-disease associations, gene expression patterns, protein-protein interaction networks, and real-world clinical data [22, 51, 66]. Techniques such as deep learning, graph neural networks, and natural language processing are employed to uncover hidden relationships and predict drug efficacy across different pathological contexts such as dementia [66, 20, 18].

Personalized medicine has become one of the most significant advances in the treatment of dementia diseases, and artificial intelligence plays an essential role in this approach. Instead of applying standard treatments to all patients, personalised medicine seeks to adapt treatments to the individual characteristics of each person, taking into account their genetics, medical history and lifestyle. In this context, artificial intelligence is key to analyzing and managing large volumes of data, enabling more precise and effective therapeutic recommendations. One of the main advances in AI-driven personalized medicine is the analysis of genetic data [108, 92, 63]. Each patient exhibits unique genetic variability that can influence their response to treatments. Although personalized treatment strategies for dementia diseases like dementia’s show promise, they face challenges related to both efficacy and side effects, with insufficient therapeutic effectiveness often representing the greater hurdle. Artificial intelligence algorithms are capable of analyzing large-scale genetic variations and detecting complex patterns that may not be easily identified by clinicians. While this enables a more tailored approach to treatment, the challenge remains that current therapies often lack sufficient efficacy in slowing disease progression, despite personalized adjustments based on genetic factors. In this way, the selection of treatments is optimised, reducing the likelihood of side effects and increasing therapeutic efficacy [43]. In addition to genetic data, these systems also consider clinical and lifestyle factors in their analysis [71]. Electronic medical records, brain images, and data on the patient’s physical and mental activity are integrated to provide a more holistic view of their health. This allows algorithms to design fully personalized treatment plans tailored to each patient’s specific needs, such as IBM Watson Health, Google DeepMind Health, Philips HealthSuite or PathAI among others.

## 5. Challenges and Ethical Considerations

The integration of artificial intelligence in the diagnosis, monitoring, and treatment of dementia diseases brings with it numerous challenges, particularly regarding data privacy, clinical validation, and ethical concerns. These challenges must be addressed to ensure the responsible deployment of AI technologies in healthcare in compliance with regulatory frameworks, including European, American, and international standards.

### 5.1. Data Privacy and Security

One of the primary challenges in applying artificial intelligence techniques in healthcare is ensuring the privacy and security of patient data. The use of AI in managing dementia diseases often involves collecting and analyzing large, sensitive datasets, including medical imaging, biomarker information, genetic data, and personal health records. Given the inherently sensitive nature of these datasets, safeguarding patient information is paramount. To address these privacy concerns, innovative approaches such as federated learning have been proposed [48]. These algorithms allow AI models to be trained without transferring sensitive data to a central server, keeping the data at its original location, whether in hospitals, clinics, or other healthcare facilities. This decentralized approach ensures that patient data never leaves local systems, thus minimizing the risk of data breaches. By enabling distributed collaboration among multiple institutions, federated learning facilitates the development of AI models without compromising patient privacy [77].

Furthermore, AI-supported data anonymization techniques are critical in ensuring that processed data cannot be traced back to individuals. Anonymization, which involves the removal or transformation of identifying information, ensures that datasets can be safely used for research and AI model development without violating patient privacy. This process mitigates the risks associated with handling sensitive data, allowing for its secure use in medical applications. In the European Union, the General Data Protection Regulation (GDPR) provides a comprehensive legal framework for the protection of personal data, ensuring that patient information is processed securely and transparently [2]. The GDPR mandates stringent consent requirements, which emphasize that individuals must be fully informed about how their data will be used. It also ensures that patients retain the right to access, correct, and delete their data. Moreover, the regulation imposes strict controls over data storage and transfer, particularly when sensitive information is processed by AI systems, thereby reinforcing the importance of maintaining data security throughout the research and application phases. Complementing the GDPR, the Artificial Intelligence Act (AI Act) [4], recently approved by the European Union, establishes a risk-based regulatory framework for AI systems. It introduces specific obligations for high-risk AI applications, including those in healthcare, requiring robust data governance measures, transparency, and accountability. The AI Act mandates rigorous testing, documentation, and human oversight of AI tools to ensure that they are trustworthy, safe, and aligned with fundamental rights, including the right to privacy. This regulation is particularly relevant when deploying AI models that interact with patient data, as it strengthens legal safeguards and harmonizes ethical standards across the EU.

In the United States, the Health Insurance Portability and Accountability Act (HIPAA) sets standards for the protection of health information [1]. HIPAA governs the use and disclosure of Protected Health Information (PHI) and establishes security protocols to ensure that patient data is protected, both during transmission and while stored. However, given the rapid pace of AI development and the increasing use of health data by third-party companies, the evolving landscape of data privacy laws may require further adaptations to address emerging concerns related to AI applications. Beyond these regional regulations, international standards such as those set by the International Organization for Standardization (ISO) [3] and the World Health Organization (WHO) [103] emphasize the need for secure, interoperable, and ethical data-handling practices, ensuring that AI systems respect privacy while enabling effective healthcare solutions.

### 5.2. Clinical Validation and Trust

For AI-based solutions to be widely adopted in clinical practice, they must undergo rigorous validation to ensure their safety and efficacy [65]. Although AI models show great promise for the diagnosis and treatment of dementia diseases, these systems must be tested with robust and diverse data sets that reflect the heterogeneity of the patient population. Clinical trials designed to validate AI tools must comply with international standards, such as the guidelines of the International Council for Harmonisation of Technical Requirements for Pharmaceuticals for Human Use (ICH), to ensure that these models work reliably in real-world settings [68]. In addition, it is necessary to take into account the AI-reliability to be accepted by both doctors and patients [8]. AI systems must be transparent in their decision-making processes, allowing healthcare professionals to understand how decisions are made and on what basis. Explainable AI (XAI), is a growing area of research that aims to make machine learning models more interpretable, which is crucial for building trust among clinicians who rely on AI tools for critical decision-making. Within XAI, several techniques improve the interpretability of models in the detection and monitoring of dementia diseases [6, 7].

Several XAI techniques can be employed to increase transparency and interpretability in AI models for dementia diseases. These techniques can be broadly categorized based on the type of data they use and how they help clinicians understand model predictions. For instance, SHAP (SHapley Additive Explanations) and LIME (Local Interpretable Model-agnostic Explanations) are feature attribution methods that reveal the contribution of each data feature in the model’s decision-making process. SHAP uses Shapley values to quantify the contribution of each feature in any type of data, including brain images and clinical measurements, and is particularly useful for identifying key biomarkers influencing diagnosis, such as brain atrophy or cognitive test scores. LIME generates local perturbations in input data to explain specific predictions, which is particularly useful for providing local explanations for AI-based classifications of brain scans or physiological signals [46].

In addition, Grad-CAM (Gradient-weighted Class Activation Mapping) and saliency maps are visualization-based techniques that help clinicians understand the most influential image regions in the model’s prediction. Grad-CAM uses gradients from deep networks to create heat maps over images, highlighting critical brain regions in MRI or PET scans for early disease detection. Saliency maps follow a similar approach, but focus on the individual pixels that most influence the output, which is useful for detecting structural brain changes contributing to AI predictions [86, 46].

Other approaches such as decision trees and random forests offer explicit, rule-based models for classifying patients based on clinical data, making the model’s logic more transparent. These techniques provide rule-based, interpretable predictions for disease classification, which are helpful for clinicians when making decisions based on a patient’s clinical history and test results. Furthermore, generalized additive models (GAMs) and explainable boosting machines (EBMs) provide a balance between accuracy and interpretability, modeling the interaction between variables while keeping the decision-making process understandable. These models are particularly useful in predicting disease progression with interpretable decision-making [12, 81]. Lastly, counterfactual explanations enable clinicians to see how changes in a patient’s characteristics—such as hippocampal volume or cognitive test scores—would impact the model’s prediction, offering insights into how modifying patient features would affect diagnosis and disease progression [96, 64]. By employing these XAI techniques, it is possible to provide a clearer and more interpretable rationale for clinicians, increasing their confidence in these tools for patient care.

### 5.3. Ethical Considerations

The application of AI in healthcare also raises several ethical concerns, particularly related to autonomy, fairness, and accountability. One of the core ethical issues is ensuring that AI tools do not inadvertently reinforce existing biases in healthcare. If AI models are trained on biased datasets, they may produce skewed results that disproportionately affect certain patient populations, particularly vulnerable or underrepresented groups. This can lead to health disparities and exacerbate existing inequalities in healthcare access and outcomes. To mitigate this, it is essential to ensure that training data is diverse and representative of the broad range of patients who are likely to benefit from AI-driven solutions.

Additionally, regulatory bodies such as the European Medicines Agency (EMA) and the U.S. Food and Drug Administration (FDA) are placing increased emphasis on fairness in AI applications, encouraging developers to implement strategies for identifying and addressing bias within their algorithms. Another ethical consideration is the issue of informed consent. AI tools that provide diagnostic or treatment recommendations must ensure that patients understand the role of AI in their healthcare and consent to its use. Clear communication about the limitations of AI, the potential risks involved, and the role of human oversight is essential for ensuring that patients can make informed decisions regarding their treatment. Furthermore, the increasing reliance on AI in healthcare raises questions of accountability. When AI systems are involved in clinical decision-making, it is critical to determine who is ultimately responsible for the outcomes, particularly in the case of misdiagnoses or treatment failures. Establishing clear frameworks of responsibility is necessary to ensure that both healthcare providers and developers are held accountable for the performance and impact of AI systems.

The global nature of AI in healthcare necessitates international collaboration to establish unified standards for AI development and deployment. While the regulatory landscape in the EU and the US is advancing, different countries have varying requirements, which can complicate the development of these systems that can be deployed worldwide. For example, the European Union’s Medical Device Regulation (MDR) and the In Vitro Diagnostic Regulation (IVDR) offer specific guidelines for AI tools in healthcare, while the FDA in the US has its own set of regulatory pathways.

In this context, international cooperation is essential to harmonize these standards, ensuring that AI solutions meet high safety, efficacy, and ethical standards, regardless of geographical location. Collaboration between regulatory bodies, researchers, developers, and healthcare professionals will be key to overcoming challenges related to crossborder data sharing, clinical trial designs, and the ethical deployment of AI technologies.

## 6. Clinical Adoption

While numerous artificial intelligence methods have demonstrated great potential in research contexts, the transition to real-world clinical practice is not straightforward. The successful implementation of AI in clinical environments requires addressing technological and regulatory barriers, adapting to existing clinical workflows, and fostering trust among healthcare professionals. One of the main barriers is the skepticism from clinicians, who are often unfamiliar with AI technologies and may be resistant to adopting tools that they perceive as black boxes. As argued by Thibeau-Sutre et al. (2023) and Gaur et al. (2024), many of these tools do not offer interpretable outputs and cannot be easily incorporated into decision-making processes that require traceability and justification [93, 28]. Especially in fields like dementia care, clinicians often prefer interpretable, heuristic-based reasoning, and are reluctant to trust systems that do not provide clear explanations for their outputs. To overcome this, AI tools must be designed to provide not only accurate predictions but also intelligible reasoning paths. Clinicians must be able to understand why an AI model made a certain recommendation, particularly when it may influence diagnosis or treatment plans. Educational efforts and hands-on training sessions are also essential to improve clinicians’ familiarity and confidence with AI systems.

Another major challenge is the interoperability between AI tools and the diverse technological infrastructure found in hospitals. Many healthcare facilities use fragmented systems—electronic health records (EHRs), radiology imaging viewers, lab data repositories—that are not designed to easily integrate external AI tools. In the context of dementia, which often requires longitudinal monitoring and multimodal data (e.g., MRI, PET, cognitive tests), the ability of AI models to interface smoothly with these systems is crucial. Moreover, the operational workflow must be taken into account. AI tools must be seamlessly embedded into existing clinical routines to avoid workflow disruptions. For instance, if clinicians are forced to use separate interfaces or manually transfer data to and from the AI tool, adoption rates will likely be low. Designing AI systems that can be accessed through existing clinical interfaces, such as EHR dashboards or radiology viewers, is key to ensuring usability and scalability. To facilitate seamless integration, it is essential to adopt interoperability standards such as HL7 or FHIR, which enable efficient communication between AI systems and clinical platforms. From a regulatory perspective, these systems in healthcare are subject to increasing scrutiny.

Ethical concerns are also paramount. As [42] highlights, AI systems must be developed in ways that uphold patient autonomy, fairness, and privacy. In dementia care, where patients may not be able to provide informed consent, data governance and transparency become even more important. This can exacerbate existing healthcare disparities and undermine the reliability of AI-supported diagnoses. Table 7 summarizes key challenges and corresponding strategies for effective knowledge transfer and integration of AI into clinical practice while Table 6 focuses on dementia.

**Table 6.**
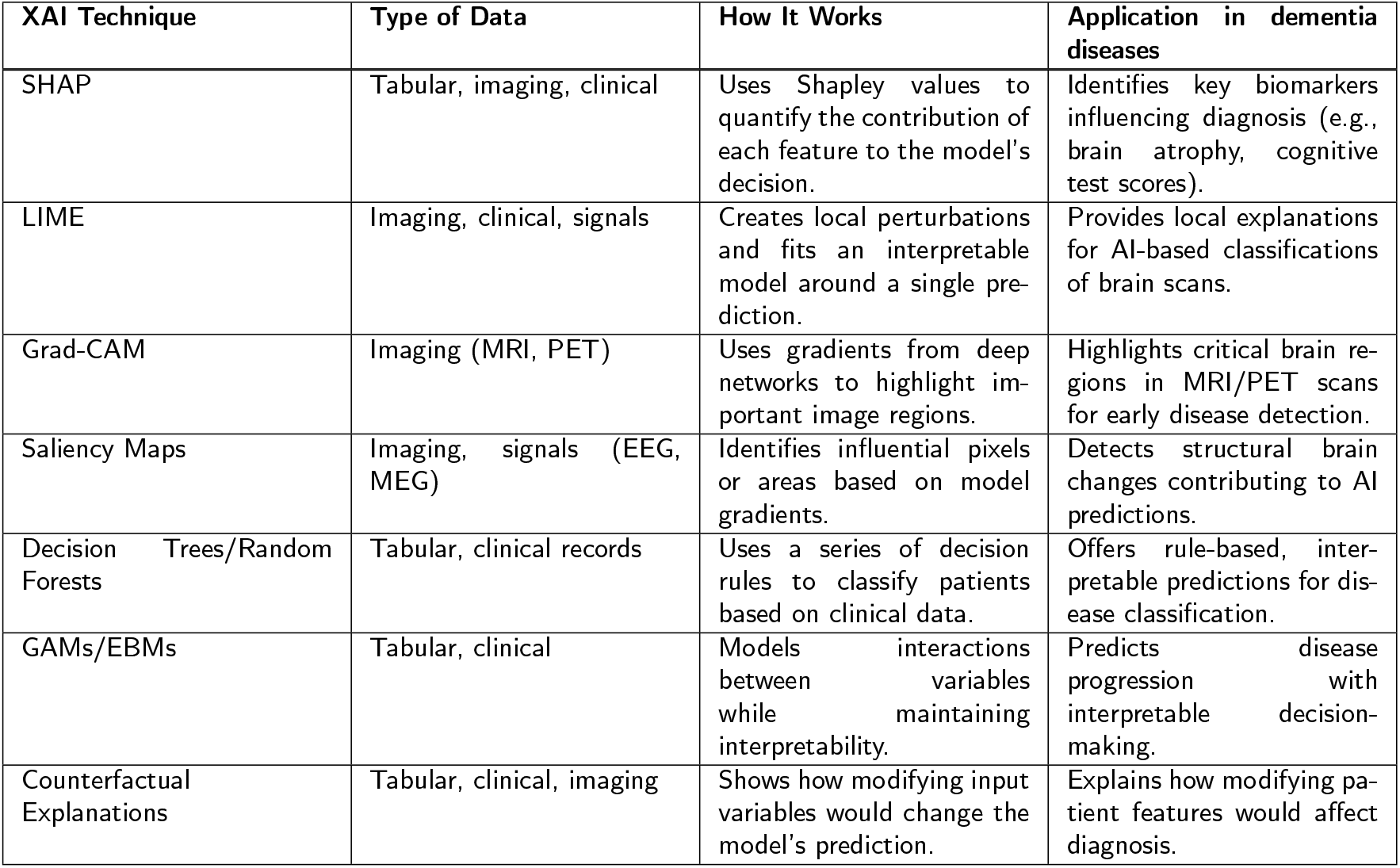
Explainable AI techniques applied to dementia diseases.

**Table 7.**
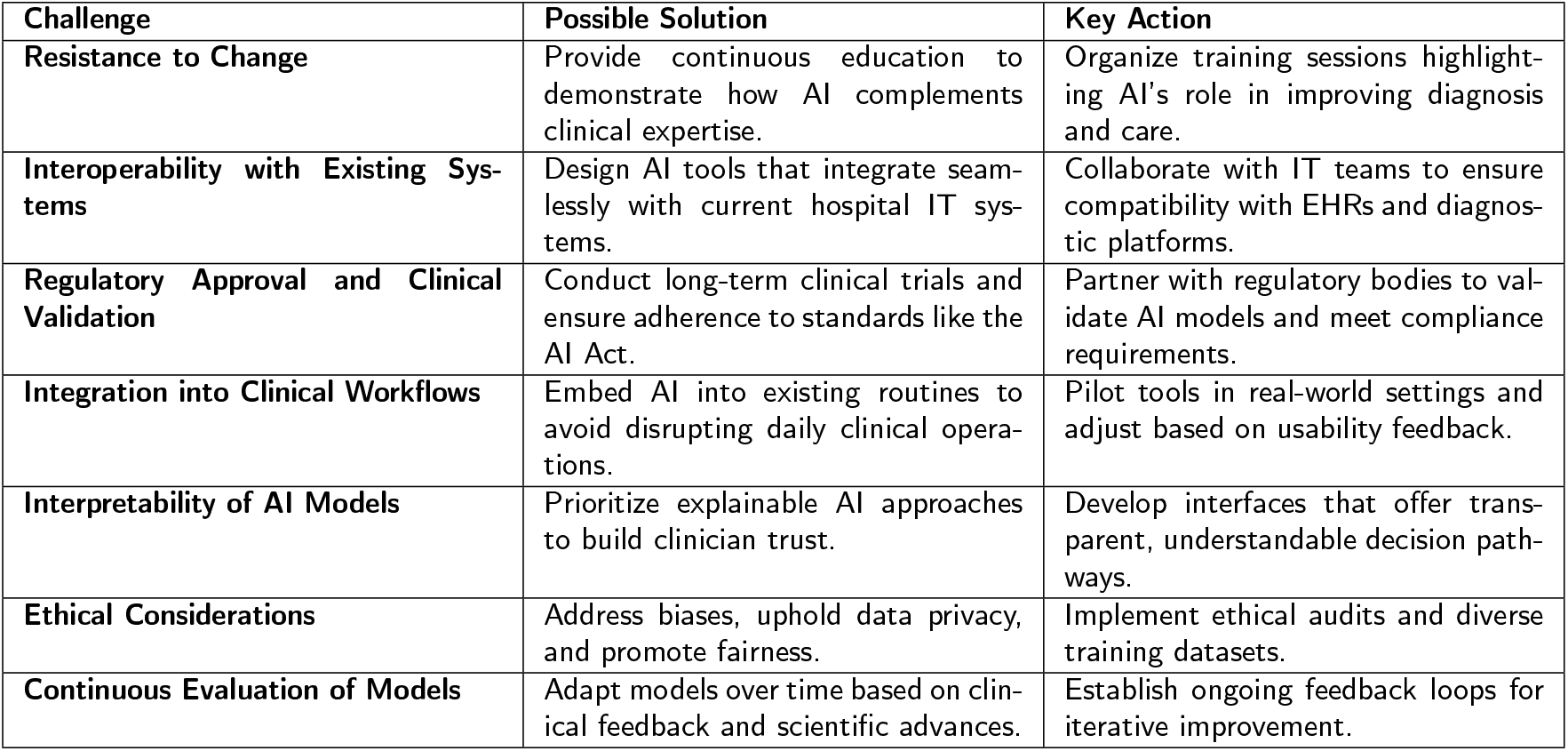
Challenges and Solutions for Integrating AI into Clinical Practice.

| Challenge | Possible Solution | Key Action |
| --- | --- | --- |
| **Resistance to Change** | Provide continuous education to demonstrate how AI complements clinical expertise. | Organize training sessions highlighting AI's role in improving diagnosis and care. |
| **Interoperability with Existing Systems** | Design AI tools that integrate seamlessly with current hospital IT systems. | Collaborate with IT teams to ensure compatibility with EHRs and diagnostic platforms. |
| **Regulatory Approval and Clinical Validation** | Conduct long-term clinical trials and ensure adherence to standards like the AI Act. | Partner with regulatory bodies to validate AI models and meet compliance requirements. |
| **Integration into Clinical Workflows** | Embed AI into existing routines to avoid disrupting daily clinical operations. | Pilot tools in real-world settings and adjust based on usability feedback. |
| **Interpretability of AI Models** | Prioritize explainable AI approaches to build clinician trust. | Develop interfaces that offer transparent, understandable decision pathways. |
| **Ethical Considerations** | Address biases, uphold data privacy, and promote fairness. | Implement ethical audits and diverse training datasets. |
| **Continuous Evaluation of Models** | Adapt models over time based on clinical feedback and scientific advances. | Establish ongoing feedback loops for iterative improvement. |

**Table 8.**
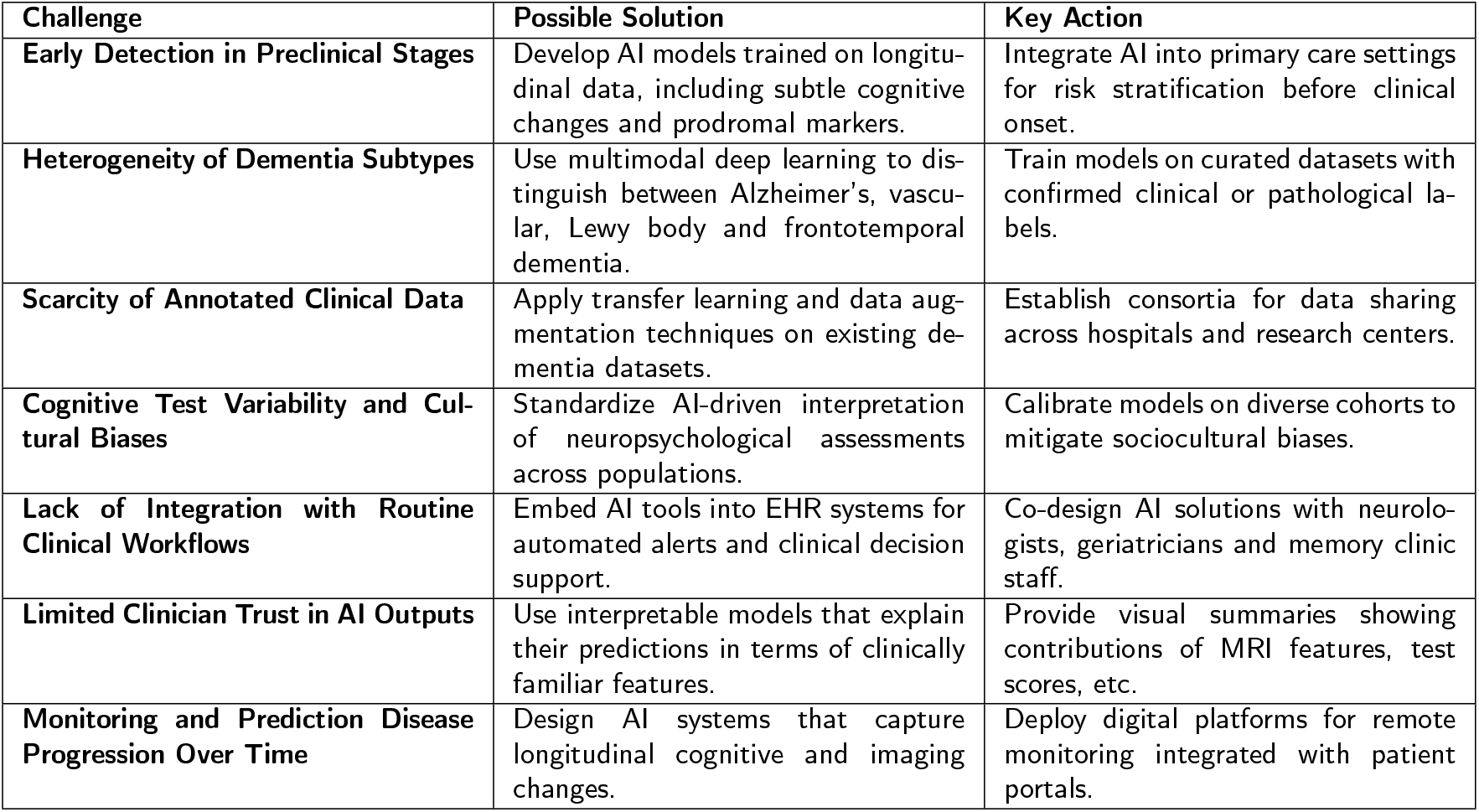
Specific Challenges and AI-Focused Solutions for Dementia Diagnosis and Monitoring.

Another critical point is the continuous evaluation of deployed AI models. In dementia, disease progression is nonlinear and varies greatly between individuals. As new clinical evidence, biomarkers, or imaging modalities become available, AI models must be updated accordingly. Without mechanisms for iterative learning and model revision, the utility of AI tools will diminish over time. Effective collaboration between clinicians, developers, and regulatory bodies is necessary to implement such feedback loops.

Finally, legal and insurance frameworks must evolve to accommodate AI. Currently, if an AI-based diagnostic tool produces an erroneous result that leads to patient harm, the question of liability remains unresolved. Most patients are not entitled to compensation when harmed by an AI-supported diagnosis. This legal gap can hinder widespread adoption and may expose clinicians to additional risk unless clear accountability structures are established.

## 7. Future Directions and Agentic AI

As the field of artificial intelligence progresses rapidly, its intersection with dementia’s disease research is poised to undergo a profound transformation. Beyond the current generation of predictive models and diagnostic tools, emerging paradigms in AI offer the potential to fundamentally reshape how data is interpreted, integrated, and acted upon in both research and clinical practice. One of the most promising frontiers is the development of agentic AI—systems that exhibit autonomous goal-setting, planning, and adaptive decision-making [45]. These models, exemplified by recent architectures such as AutoGPT, are capable of decomposing complex clinical objectives into iterative tasks, dynamically retrieving and synthesizing multimodal information, and continuously refining their outputs based on environmental feedback. In the context of dementia and focusing on Alzheimer’s disease, where longitudinal, heterogeneous data are the norm, agentic AI could support autonomous monitoring, hypothesis generation, and proactive care coordination, significantly reducing the cognitive and administrative burden on clinicians while enhancing patient outcomes [109].

Simultaneously, the rise of large-scale foundation models—particularly those trained on vast corpora of visual or textual data—offers new opportunities for transfer learning across domains. While such models have demonstrated exceptional generalization capabilities, their deployment in dementia research faces challenges due to domain mismatch and the scarcity of labeled data. Addressing these limitations through advanced domain adaptation techniques, such as domain-specific fine-tuning and hybrid transfer frameworks, may enable the leveraging of pretrained architectures to extract meaningful patterns from neuroimaging, cognitive assessments, and clinical narratives specific to AD populations. This is especially relevant in settings where high-quality annotated datasets are difficult to acquire.

To further address the limitations of low-data environments, self-supervised learning (SSL) has emerged as a compelling strategy. This technique allows models to learn rich, generalizable representations from unlabeled data by exploiting intrinsic structures, such as spatial, temporal, or semantic correlations. This is particularly valuable in dementia research, where labeled datasets are often small, imbalanced, or clinically heterogeneous [33, 54]. By incorporating SSL into pipelines for structural and functional neuroimaging, speech analysis, or digital biomarkers, it is possible to build more robust and scalable models capable of performing reliably across diverse patient cohorts and clinical settings [31]. Another critical development lies in integrating large language models (LLMs) with autonomous agents to support complex clinical reasoning and knowledge synthesis. In a disease characterized by gradual progression and multifaceted clinical histories, such integrations can facilitate the automated summarization of electronic health records over extended timelines, the contextual interpretation of diagnostic findings, and the generation of individualized care recommendations [13]. Importantly, this synergy enhances not only the analytical capacity of AI systems but also their interpretability and traceability, aligning with the growing demand for explainability, transparency, and ethical rigor in clinical AI applications. Taken together, these emerging directions signal a shift from narrow, task-specific tools toward holistic, adaptive, and context-aware AI systems. Realizing this potential will require multidisciplinary collaboration, careful validation in prospective settings, and the continuous integration of ethical, regulatory, and clinical perspectives. Nonetheless, the convergence of agentic reasoning, self-supervised learning, and large-scale foundation models represents a transformative opportunity for the future of AI in dementia.

## 8. Conclusions

This review has provided a comprehensive analysis of the role of artificial intelligence in the study, diagnosis, and management of dementia, emphasizing its transformative impact on early detection, biomarker identification, and therapeutic development. AI-driven approaches, particularly machine learning and deep learning models, have revolutionized disease modeling by integrating and analyzing large-scale multimodal datasets, including neuroimaging, genomic, proteomic, and clinical data. This capability enhances diagnostic precision, aids in disease stratification, and facilitates the development of personalized treatment strategies.

These novel techniques accelerate data interpretation and predictive modeling but also provides new computational frameworks for understanding disease mechanisms. Advanced methodologies such as transformer models and multimodal approaches have demonstrated remarkable success in neuroimaging analysis, automating the detection of structural and functional abnormalities linked to neurodegeneration. Additionally, AI-based systems have contributed to discovering novel pharmacological targets and optimizing drug repurposing strategies, expediting therapeutic advancements.

Despite these breakthroughs, significant challenges remain. Data heterogeneity, lack of standardized protocols, and biases in training datasets can compromise model generalizability and clinical applicability. Furthermore, the interpretability of AI-generated insights remains a major barrier to clinical adoption, necessitating the development of explainable AI models that provide transparent and biologically meaningful predictions. Ethical considerations, including patient privacy, informed consent, and equitable access to AI-driven healthcare solutions, must also be addressed to ensure responsible deployment.

To fully harness the potential of AI in dementia disease research, future efforts should focus on enhancing model robustness, integrating multi-omics data, and fostering interdisciplinary collaborations between neuroscientists, clinicians, and AI researchers. Regulatory frameworks and rigorous validation studies will be crucial for translating AI innovations into clinical practice. Ultimately, the continued convergence of AI and neuroscience holds immense promise for improving early diagnosis, refining treatment strategies, and advancing our mechanistic understanding of dementia disorders, bringing us closer to more effective and personalized therapeutic solutions.

## Supporting information

Data Extraction

## Data Availability

Systematic review.

## Notes

### Competing Interest Statement

The authors have declared no competing interest.

### Funding Statement

No funding

